# CLINICAL DIAGNOSIS CODES IDENTIFY PATIENTS UNLIKELY TO RECEIVE ORDERS FOR FECAL IMMUNOCHEMICAL TESTS

**DOI:** 10.1101/2025.06.07.25327375

**Authors:** Andrew Tong Li, Shohei Burns, Dalia Martinez, Patricia Wang, Sona Aggarwal, Michael Potter, Urmimala Sarkar, Ma Somsouk

## Abstract

**Introduction:** Organized screening programs automate fecal immunochemical test (FIT) outreach based on chronological age. However, providers and patients may defer screening due to coexisting conditions, which can be captured by International Classification of Disease, 10^th^ revision (ICD-10) codes. We sought to identify codes associated with not having a FIT order.

**Methods:** We included screen-eligible patients with a primary care visit between June 2022 and June 2023. We enumerated and compared the frequency of each ICD-10 code between patients with and without a FIT ordered. We conducted a subgroup analysis including only those with a Charlson comorbidity index (CCI) <5.

**Results:** We identified 15,020 screen-eligible patients, with 10,187 (67.8%) patients having had a FIT order and 4,833 (32.3%) patients without a FIT order. Of the 1,215 ICD-10 codes examined, 96 were significantly associated with not having a FIT order. One broad category of codes pertained to digestive diseases such as benign and malignant colonic neoplasms (ICD-10 code C18, odds ratio (OR) 0.06, 95% CI [0.001-0.44]) and diverticular disease (K57, OR 0.26 [0.18-0.36]). Another category was comorbid conditions which included frailty (R54, OR 0.14 [0.03-0.55]) and paralysis (G82, OR 0.14 [0.03-0.55]). Those with acute conditions such as cervical fractures (S12, OR 0.23 [0.10-0.48]) and cryptococcosis (B45, OR 0.05 [0.001-0.38]) were also less likely to have a FIT order. Among the patients with CCI<5, 41 codes were significantly associated with not having a FIT order, including heart failure (I50, OR 0.55 [0.43-0.71]) and chronic kidney disease (N18, OR 0.58 [0.43-0.77]).

**Conclusion:** Patients deferred from screening were more likely to have ICD-10 codes signifying digestive diseases, comorbidities, and acute conditions. Even among patients with fewer co-morbid conditions, we identified health conditions that were associated with screening deferral. Future work should consider whether screening programs could incorporate ICD-10 codes to align FIT outreach more closely with provider and patient preferences.

## Introduction

Colorectal cancer (CRC) screening has effectively reduced CRC incidence and mortality.^1^ In many health systems with lower rates of screening, including safety-net health systems, CRC screening has been opportunistic.^2^ Organized screening programs within health systems like Kaiser Permanente have been shown to achieve high rates of screening.^3^ CRC guidelines use chronological age to guide screening while allowing for the exclusion of patients with limited life expectancy. Meanwhile, there is no standard practice when considering the role of a patient’s health status, though the Charlson Comorbidity Index and prognostic calculators have been proposed to guide decision making regarding the risk and benefit of screening.^4^

Organized screening programs using age-based algorithms risk overscreening patients with multiple comorbidities.^5^ Ideally, screening programs that automate outreach would align outreach with the patient’s preference and a provider’s intention to offer screening. While adherence to CRC screening incorporates the provider’s intention to offer it and the patient’s willingness to complete it, an order for a service like the fecal immunochemical test (FIT) more directly reflects the provider’s intent to screen.

We postulate that structured data elements such as the International Classification of Disease – 10th Revision (ICD-10) codes can be used to capture conventional clinical diagnoses and conditions, such as limited life expectancy, that are associated with a decreased likelihood of receipt of a FIT order; in addition, we expect that uncover coexisting illnesses not traditionally linked to limited life expectancy will be associated with deferral of screening. This retrospective observational study extracted patient-level data elements from electronic health records to examine the association between individual ICD-10 codes and the FIT order status. Identification of clinical codes could then improve the delivery of population-based screening services as intended by providers.

## Methods

### Cohort Development and Definition

We included screen-eligible patients ages 50 to 75 in a safety-net health system with at least one primary care provider (PCP) encounter from June 26, 2022 to June 26, 2023 (Figure 1). During the study period, screening eligibility was based on age between 50 and 75. Although the recommended age to begin CRC screening was lowered from 50 to 45 in May 2021, the electronic medical record had not yet been updated to include care gap alerts for patients 45 to 49. We excluded patients from the analytic cohort if they had a documented colonoscopy in the past 10 years or if they ever had a positive FIT result at any time, since these patients would also be ineligible for FIT screening Screen-eligible patients were categorized as either having a FIT ordered or no FIT ordered between June 26, 2022, and June 26, 2023. In this safety-net health system, FIT is a commonly offered form of CRC screening, while colonoscopy can be selected at the discretion of either the PCP or the patient. Since colonoscopy is performed for both screening and diagnostic indications, and the goal of the analysis is to understand FIT ordering patterns, we did not use colonoscopy to define the cohorts.

**Figure 1.**
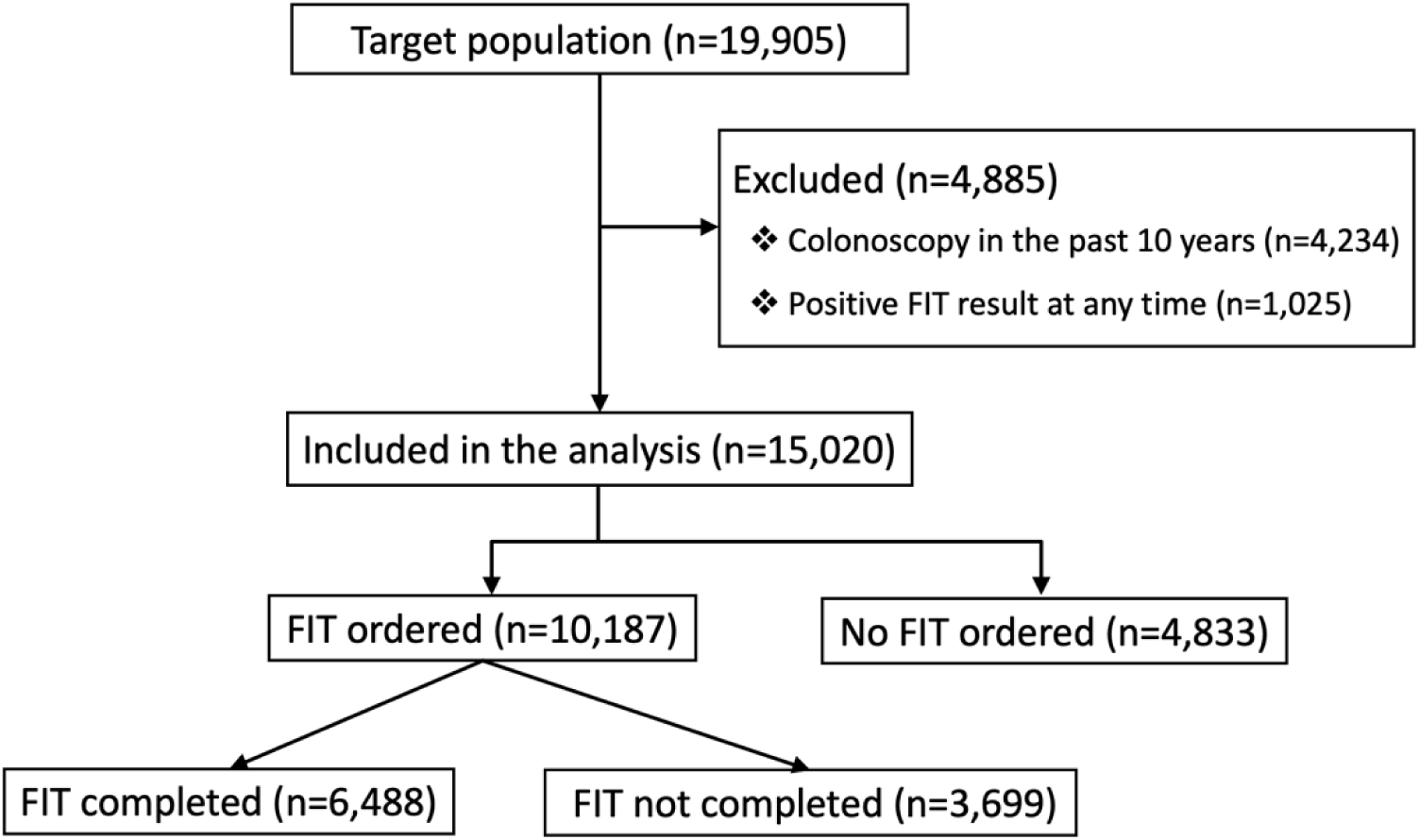
Flow Diagram of the patients eligible for CRC screening with a primary care encounter. Patients included in the analysis were divided by the occurrence of a FIT order.

### Covariates and outcome variable

In addition to patient demographic data, we extracted all orders for FIT and associated ICD-10 codes and the Charlson Comorbidity Index (CCI) for each eligible patient. All ICD-10 codes were collapsed into the 3 positions that precede the decimal (Figure 2) to aggregate clinical conditions into similar categories. For instance, M1A.312 (chronic gout of the wrist) and M1A.432 (chronic gout with renal impairment) would both be collapsed to M1A (chronic gout).

**Figure 2.**
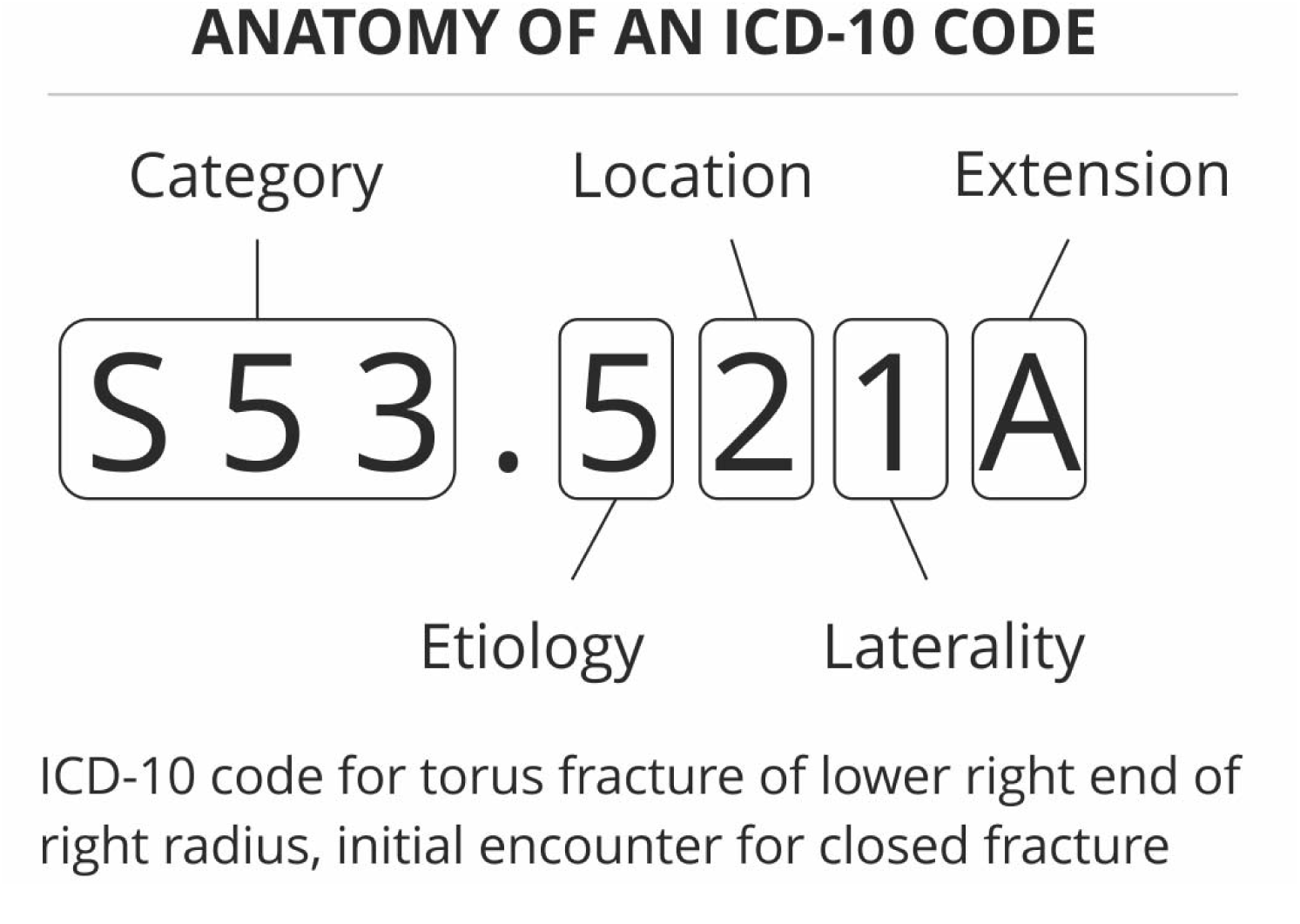
Anatomy of an ICD-10 code. The 3 characters preceding the decimal is the collapsed category code. Reproduced from https://blogs.halodoc.io/automatic-icd-10-code-assignment-to-consultations-using-deep-learning/.

### Analysis

We summarized patient demographic characteristics for the two cohorts, “Patients with FIT ordered” and “Patients with no FIT ordered,” using proportions and means as appropriate. The primary analysis enumerated and compared the frequency of each ICD-10 code between the two cohorts using the odds ratios (OR) along with the corresponding 95% confidence interval (CI) using the Fisher’s exact test with a Benjamini-Hochberg correction for multiple hypothesis testing.^6^ Given the exploratory nature of this study, we focused our attention not only on statistical significance, but also on substantially low odds ratios set at a threshold of less than 0.6, which were presumed to be more clinically meaningful.

A secondary analysis was performed among patients who had fewer documented comorbidities, as measured by a CCI less than 5. Since screening guidelines state that individuals with limited life expectancy are unlikely to benefit from screening, we hypothesized that among these patients, factors besides life expectancy would be associated with deferred screening. Therefore, we repeated the analysis on a restricted cohort of patients with CCI lower than 5 in order to examine if ICD-10 codes could identify coexisting conditions that were associated with deferred screening.

All statistical analyses, figures, and tables were done with Microsoft Excel (version 16), RStudio (version 4.3.1), and Stata MP 18.

## Results

### Patient Demographics

There was a total of 19,905 patients aged 50-75 in the health network with a primary care encounter in the study period, and a total of 15,020 patients were eligible for CRC screening. Of the patients eligible for screening, 10,187 (67.8%) patients had a FIT order (FIT ordered group), and 4,833 (32.3%) patients did not have a FIT order (No FIT ordered group) (Table 1). In the FIT-ordered group, 6,488 (63.7%) completed their FIT. Overall, in the “No FIT ordered” group, the patients tended to be older, more likely to be African American, English-speaking, Medicare recipients, divorced, and unhoused.

**Table 1.** Demographic characteristics for patients eligible for CRC screening and the proportion of patients with and without an order for FIT.

| Category | Total | Cohort |  |
| --- | --- | --- | --- |
|  |  | Patients with FIT ordered | Patients with no FIT ordered |
| Patient Count | 15,020 (100%) | 10,187 (67.8%) | 4,833 (32.2%) |
| **Sex, n (%)** |  |  |  |
| Female | 7,632 (100%) | 5,385 (70.6%) | 2,247 (29.4%) |
| Male | 7,379 (100%) | 4,797 (65%) | 2,582 (35%) |
| Other/Unknown | 9 (100%) | 5 (55.6%) | 4 (44.4%) |
| **Age, n (%)** |  |  |  |
| 50-54 | 2,965 (100%) | 2,209 (74.5%) | 756 (25.5%) |
| 55-59 | 3,369 (100%) | 2,293 (68.1%) | 1,076 (31.9%) |
| 60-64 | 3,611 (100%) | 2,452 (67.9%) | 1,159 (32.1%) |
| 65-69 | 2,889 (100%) | 1,879 (65%) | 1,010 (35%) |
| 70-75 | 2,186 (100%) | 1,354 (61.9%) | 832 (38.1%) |
| **Race, n (%)** |  |  |  |
| Asian | 5,121 (100%) | 3,686 (72%) | 1,435 (28%) |
| Black Or African American | 1,988 (100%) | 1,225 (61.6%) | 763 (38.4%) |
| Hispanic or Latino | 4,220 (100%) | 2,920 (69.2%) | 1,300 (30.8%) |
| Other/Unknown | 845 (100%) | 576 (68.2%) | 269 (31.8%) |
| White | 2,846 (100%) | 1,780 (62.5%) | 1,066 (37.5%) |
| **Language, n (%)** |  |  |  |
| Chinese | 3,358 (100%) | 2,502 (74.5%) | 856 (25.5%) |
| English | 7,189 (100%) | 4,495 (62.5%) | 2,694 (37.5%) |
| Other/Unknown | 1,260 (100%) | 894 (71%) | 366 (29%) |
| Spanish | 3,213 (100%) | 2,296 (71.5%) | 917 (28.5%) |
| **Insurance, n (%)** |  |  |  |
| County Sponsored | 552 (100%) | 428 (77.5%) | 124 (22.5%) |
| Healthy Worker | 3,389 (100%) | 2,478 (73.1%) | 911 (26.9%) |
| Medicaid | 7,347 (100%) | 5,038 (68.6%) | 2,309 (31.4%) |
| Medicare | 3,534 (100%) | 2,118 (59.9%) | 1,416 (40.1%) |
| Other | 198 (100%) | 125 (63.1%) | 73 (36.9%) |
| **Marital Status, n (%)** |  |  |  |
| Divorced | 1,378 (100%) | 898 (65.2%) | 480 (34.8%) |
| Married | 5,253 (100%) | 3,779 (71.9%) | 1,474 (28.1%) |
| Other/Unknown | 135 (100%) | 99 (73.3%) | 36 (26.7%) |
| Separated | 572 (100%) | 379 (66.3%) | 193 (33.7%) |
| Single | 6,908 (100%) | 4,512 (65.3%) | 2,396 (34.7%) |
| Widowed | 774 (100%) | 520 (67.2%) | 254 (32.8%) |
| **Unhoused, n (%)** |  |  |  |
| Yes | 12,775 (100%) | 8,847 (69.3%) | 3,928 (30.7%) |
| No | 2,245 (100%) | 1,340 (59.7%) | 905 (40.3%) |

### ICD-10 coding categories associated with orders for FIT

We examined 1,215 ICD-10 codes. Of the 103 codes found to be significantly associated with reduced odds of FIT order, 62 (64.6%) had an OR < 0.6 and 25 (26.0%) had an OR < 0.3 (Figure 3). Conversely, 7 codes were significantly associated with an elevated likelihood of a FIT order (data not shown). Several categories of codes emerged, such as codes related to digestive disease, comorbid conditions, and acute conditions.

**Figure. 3.**
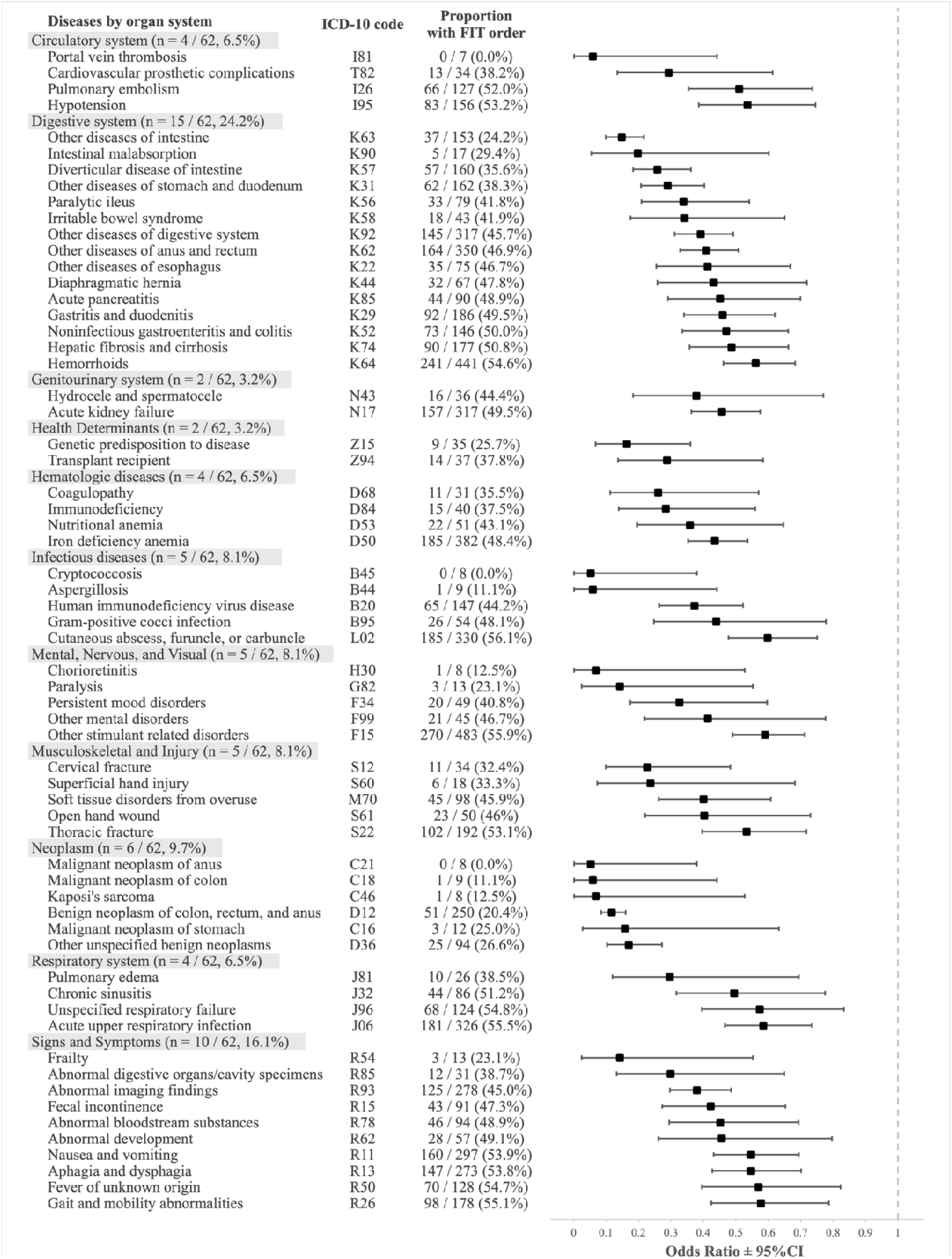
ICD-10 codes with an OR < 0.6 (n = 62), grouped by disease category, that were significantly associated with patients who did not have an order for FIT. Significance was determined to be *P* < 0.05 based on two-sided Fisher’s exact tests. Benjamini−Hochberg correction for multiple tests was applied. Codes within each category are ordered based on OR from lowest to highest.

Digestive disease codes representing benign and malignant neoplasms of the gastrointestinal tract were strongly associated with an absence of FIT orders, such as malignant neoplasm of the anus (ICD-10 code C21, odds ratio (OR) 0.05, 95% CI [0.001-0.38]), malignant neoplasm of the colon (C18, OR 0.06 [0.001-0.44]), Kaposi’s sarcoma (C46, OR 0.07 [0.002-0.53]), benign neoplasm of the colon, rectum, and anus (D12, OR 0.12 [0.08-0.16]), malignant neoplasm of the stomach (C16, OR 0.16 [0.03-0.63]), and other unspecified benign neoplasms (D36, OR 0.17 [0.10-0.27]). In addition, non-neoplastic conditions of the gastrointestinal tract associated with absence of FIT orders included intestinal malabsorption (K90, OR 0.20, 95% CI [0.05-0.60]), diverticular disease of the intestine (K57, OR 0.26 [0.18-0.36]), and other diseases of the intestine (K63, OR 0.15 [0.10-0.22]) and of the stomach and duodenum (K31, OR 0.29 [0.21-0.40]).

Comorbid conditions were also significantly associated with the absence of a FIT order. The most differentially represented clinical codes were frailty (R54, OR 0.14 [0.03-0.55]), paralysis (G82, OR 0.14 [0.03-0.55]), immunodeficiency (D84, OR 0.28 [0.14-0.56]), genetic predisposition to disease (Z15, OR 0.16 [0.07-0.36]), transplant recipient (Z94, OR 0.29 [0.14-0.58]), coagulopathy (D68, OR 0.26 [0.11-0.57]), portal vein thrombosis (I81, OR 0.06 [0.001-0.44]), and cardiovascular prosthetic complications (T82, OR 0.29 [0.13-0.61]). Other clinical conditions with an OR > 0.3 included chronic obstructive pulmonary disease (J44, OR 0.72 [0.59-0.88]), chronic kidney disease (N18, OR 0.66 [0.56-0.78]), human immunodeficiency virus disease (B20, OR 0.37 [0.26-0.52]), and heart failure (I50, OR 0.66 [0.56-0.78]).

We found that several acute clinical conditions were also associated with an absence of a FIT order. The most discriminating clinical codes with an OR < 0.3 included cervical fracture (S12, OR 0.23 [0.10-0.48]), superficial hand injury (S60, OR 0.24 [0.07-0.68]), chorioretinitis (H30, OR 0.07 [0.002-0.53], and infections such as cryptococcosis (B45, OR 0.05 [0.001-0.38]) and aspergillosis (B44, OR 0.06 [0.001-0.44]).

Of note, there were several codes associated with an elevated likelihood of having a FIT order; these included malignant neoplasm of thyroid gland (C73, OR 10.93 [1.77-449.68]), abnormal specimens from female genital organs (R87, OR 1.54 [1.17-2.04]), synovitis and tenosynovitis (M65, OR 1.37 [1.11-1.7]), abnormal blood-pressure reading (R03, OR 1.27 [1.08-1.49]), disorders of lipoprotein metabolism and other lipidemias (E78, OR 1.25 [1.14-1.37]), essential hypertension (I10, OR 1.17 [1.06-1.29]), and elevated blood glucose level (R73, OR 1.17 [1.07-1.29]).

Of the 62 ICD-10 codes with OR < 0.6, the most prevalent conditions among the 4,833 patients with no FIT ordered were other stimulant related disorders (F15, OR 0.59 [0.49-0.71]), hemorrhoids and perianal venous thrombosis (K64, OR 0.56 [0.46-0.68]), benign neoplasm of colon, rectum, and anus (D12, OR 0.12 [0.08-0.16]), iron deficiency anemia (D50, OR 0.44 [0.35-0.54]), and other diseases of anus and rectum (K62, OR 0.41 [0.33-0.51]) (Figure 4).

**Figure. 4.**
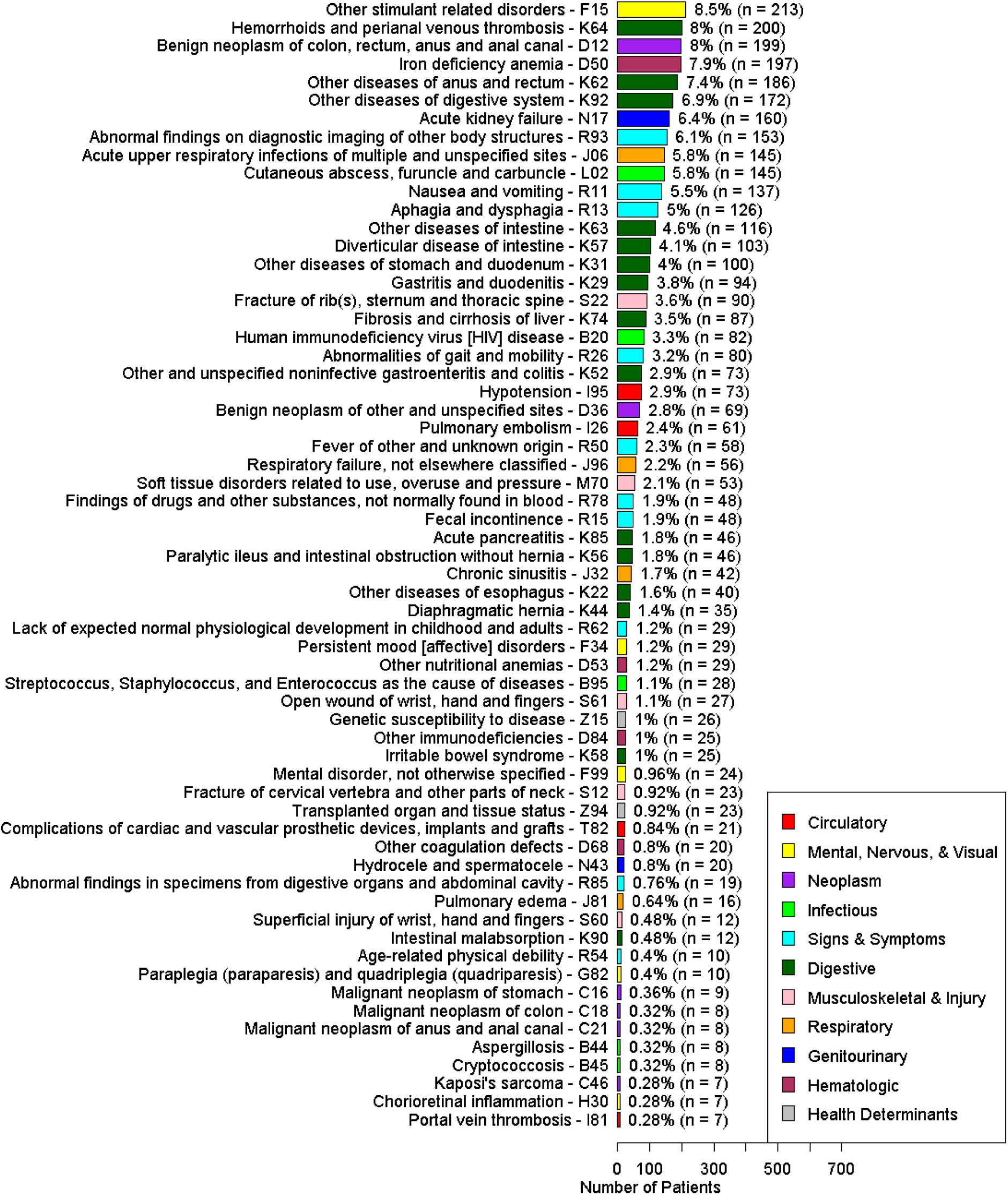
Count and proportion of patients in the cohort with no FIT ordered according to each ICD-10 code.

### ICD-10 coding categories in patients with low CCI

We found that patients with increasing comorbidities according to the CCI index were less likely to have a FIT order (74.0% CCI 0, 72.2% CCI 1, 68.5% CCI 2, 65.0% CCI 3, 64.3% CCI 4, 53.7% CCI 5+, p < 0.001). (Supplementary Figure 1). After restricting the patient population into CCI < 5 (n=12,214), there were 8,681 (71.1%) patients who had an order for FIT and 3,533 (28.9%) without an order for FIT.

There were 41 ICD-10 codes that were significantly associated with reduced odds of FIT order. There were several codes that were new or became more prominent in this analysis; they included comorbid conditions such as cachexia (R64, OR 0.09 [0.01-0.44]), heart failure (I50, OR 0.55 [0.43-0.71]), chronic kidney disease (N18, OR 0.58 [0.43-0.77]), osteomyelitis (M86, OR 0.42 [0.24-0.73]), and housing and economic circumstances (Z59, OR 0.58 [0.49-0.68]). The remaining codes are found in the supplementary file.

## Discussion

In this large, diverse population within a safety-net health system, we found that despite an encounter with the primary care clinic, over 30% of patients who were not up-to-date with CRC screening did not receive an order for a FIT test. The absence of a FIT order was significantly associated with specific patterns of clinical diagnoses and disorders, as represented by ICD-10 codes. Our analysis supports the notion that codes representing the patient’s condition factor into shared decision making and risk-benefit analysis of screening, consciously or subconsciously, resulting in an order or not for a FIT test. In this era of digitized health, the implementation of such codes in automated outreach screening programs should be studied with the broad goal of aligning services more closely with patient and provider preferences.

Unsurprisingly, we found that patients with digestive diseases and related conditions were less likely to receive a FIT order, partly because they may require a diagnostic colonoscopy and are referred accordingly.^8^ Comorbid conditions were also associated with the absence of a FIT order, such as frailty, paralysis, cirrhosis, kidney disease, obstructive pulmonary disease, heart failure, dementia, AIDS, and genetic susceptibility to disease, consistent with the practice of deferring screening in patients with lower life expectancy,^9^ the lower uptake of CRC screening demonstrated in other studies,^10–12^ and as suggested in patients with limited life expectancy.^13–15^ In contrast, milder health conditions not necessarily associated with end-organ damage, such as dyslipidemia, hyperglycemia, and hypertension, were associated with a higher rate of FIT order, consistent with previous evidence that having these health conditions increased contact with PCP which lead to more opportunities to offer screening.^11,13^

Less intuitively, we found that patients who harbored clinical codes representing acute illnesses were less likely to have an order for FIT. These conditions were related to traumatic injury, inflammation, infections, and other acute illnesses, some of which have previously been described.^11^ We additionally found that acute illnesses of the blood and circulatory system were associated with the absence of a FIT order, including portal vein thrombosis and surgical complications. This likely represents the patient’s and provider’s tacit decision to prioritize the most urgent medical issues and defer preventative care items like FIT screening until the patient’s health has stabilized.

In the population with fewer comorbidities as defined by CCI < 5, we continued to find codes for comorbid conditions such as heart failure and chronic kidney disease.

Additional codes associated with the absence of a FIT order that were not significant in the initial analysis included housing and financial problems, cachexia, and osteomyelitis. To date, no studies have published results from similar analyses that were restricted to patients with low comorbidity status as defined by CCI.

Our results suggest that providers and patients are selectively pursuing FIT. Patients with advanced comorbidities and active health conditions such as musculoskeletal fractures are not getting CRC screening. Although current guidelines discourage screening patients with limited life expectancy, life expectancy alone does not fully account for the patients’ and providers’ decisions around FIT screening. Competing medical demands, such as the need for rehabilitation following an acute injury, likely prompt patients and providers to defer FIT until the patient’s health has stabilized. As such, implementing organized screening programs that incorporate structured data elements such as ICD-10 codes as a proxy for a patient’s acute and chronic comorbid diagnoses could better align automated invitations with provider preferences for screening for each patient. These developments could pave the way for greater adoption of organized screening programs such as mailed FIT, especially in safety-net systems where acute and comorbid conditions may be more prevalent. Future studies should gather patient and provider input to better understand the rationale behind decisions to defer screening so as to inform the implementation of more targeted screening programs.

There are important limitations to this study. First, there are multiple ICD-10 codes (K63 –other diseases of intestine, D84 – other immunodeficiencies) that encompass a range of diagnoses, and while associated with FIT orders, are in and of themselves non-specific and do not provide sufficient logic as to why FIT orders were deferred. Chart review and provider interview will help refine our understanding of the provider’s intention to offer or defer screening. Second, although we used Benjamini-Hochberg correction to adjust for multiple testing and set the threshold for clinical significance at OR < 0.6, we did not perform an adjusted analysis, so spurious associations are possible. Again, qualitative studies will be needed to refine our understanding of their associations. Third, orders for FIT could have been independently made by medical assistants and nurses even if screening was not intended by the PCP. As such, inadvertent orders for FIT may have occurred among patients with these clinical codes, and if so, the misclassification of FIT orders likely underestimates the true association. We acknowledge that PCPs may not order a FIT test for other reasons, such as prior refusals by the patient, unsuccessful attempts to screen, structural constraints of the visit, such as lack of time or nursing support, failure to remember to offer a FIT, and lack of knowledge or interest around CRC screening.^16^ Moreover, there could be unconscious biases that play into the decision about when and whether a FIT test is appropriate to order, such as having different subjective judgements about the patient’s life expectancy or compliance with screening. Despite these limitations, ICD codes are available in the electronic medical record and may be explored as a valuable way to improve screening services.

In summary, we found that certain medical conditions, as captured by ICD-10 codes, were associated with a reduced likelihood of having a FIT ordered by the PCP. Current implementations of screening guidelines do not sufficiently factor in patient comorbidity, which can lead to overscreening. Advances in the way we incorporate structured data elements from the EMR could serve to streamline organized colorectal cancer screening programs, which would promote high-value care by avoiding unnecessary tests and incorporating patient preferences into medical decision-making.

## Data Availability

All data produced in the present study are contained in the manuscript. Additional raw data are available upon reasonable request to the authors.

## DISCLAIMER

The findings and conclusions in this report are those of the authors and do not necessarily represent the official position of the NIH.

## Specific author contributions

Study concept and design: MS. Acquisition of data: MS, AL, DM. Statistical analysis: MS, AL. Drafting of the manuscript: AL. Critical revision of the manuscript for important intellectual content: MS, AL, SB. Approval of the final manuscript: All authors. Study supervision: MS.

## Acknowledgements

This study would not have been possible without the support of the San Francisco Health Network of the San Francisco Department of Public Health and the UCSF Academic Research Services.

---

**Supplementary Figure. 1.**
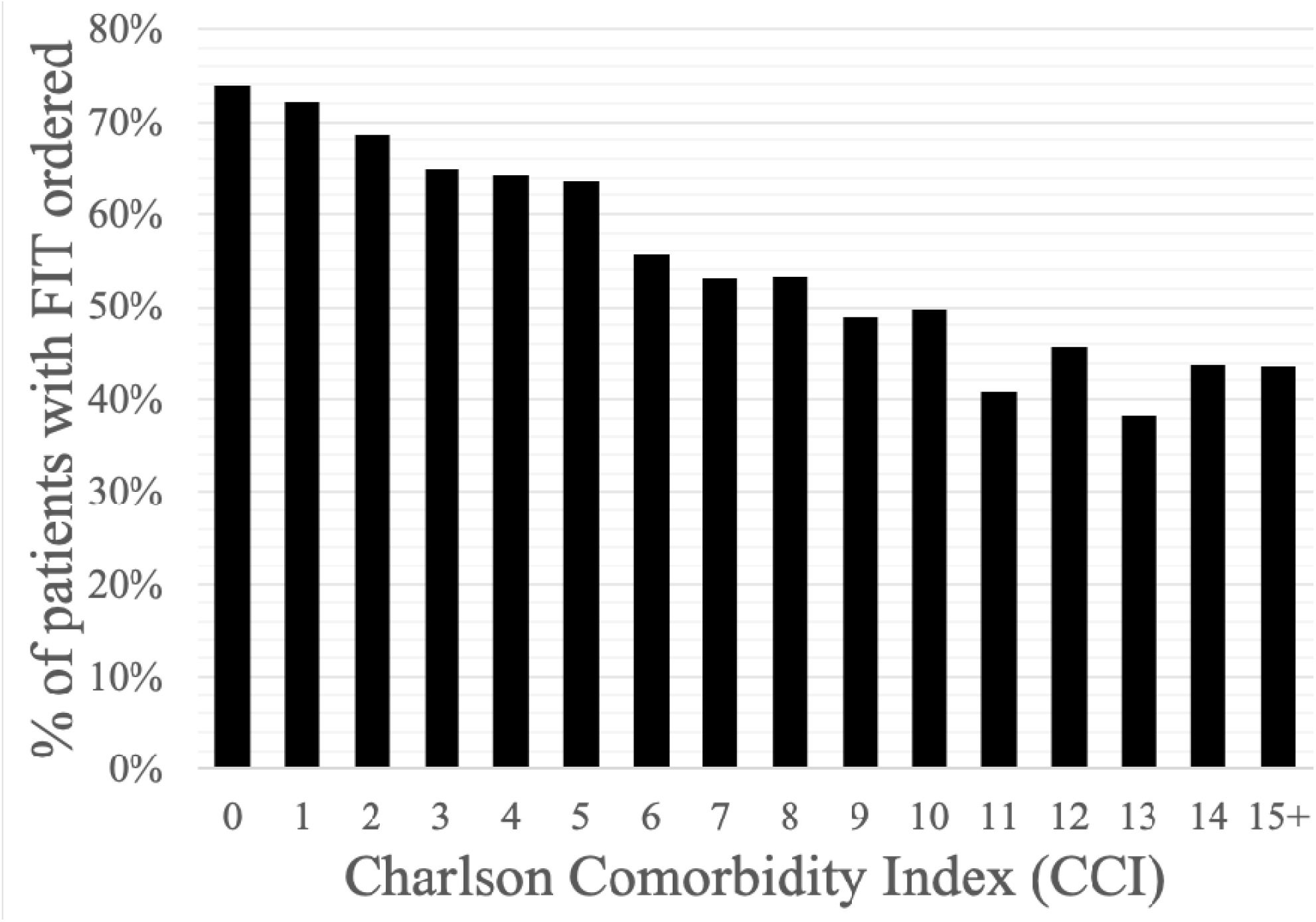
Count and proportion of patients in the cohort with FIT ordered according to Charlson Comorbidity Index (CCI).

**Supplementary Figure 2.**
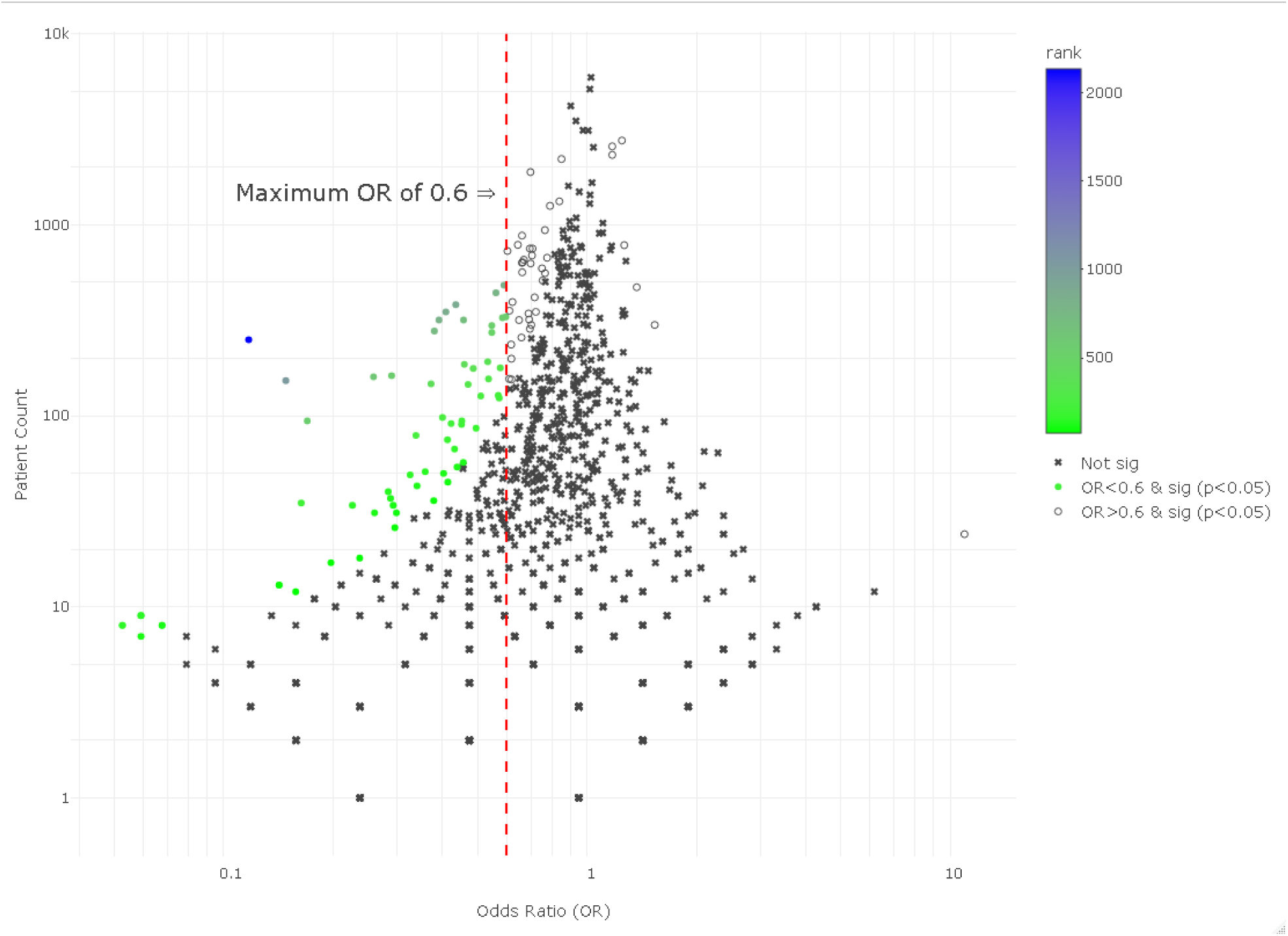
ICD-10 codes (n = 1,2*15*) distributed according to their odds ratio and patient count, plotted on a logarithmic scale. Codes with an odds ratio less than 0.6 are represented by the vertical red dashed line. *C*odes marked with an “x” *were not significant* (n = *1112*), codes with hollow circles were significant *and OR > 0.6* (n = *41*), and solid circles were significant *and OR < 0.6* (n = 6*2*). *Significance was determined to be* P *< 0.05* based on two-sided Fisher’s exact tests. Benjamini−Hochberg correction for multiple tests was applied. Rank is the ratio of patient count to odds ratio, such that high-ranked codes have a low odds ratio and high patient count.

